# High quality analysis of circulating biomarkers reveals no evidence of elevated inflammatory markers in a long COVID cohort recruited at a primary care center

**DOI:** 10.64898/2026.07.28.26359102

**Authors:** María Luz Torres, David Lerma-Irureta, Jesús Ibañez-Ruiz, Alexandre Lucas, Rosa Magallón-Botaya, Jon Schoorlemmer

**Affiliations:** Aragonese Primary Care Research Group (GAIAP), Institute for Health Research Aragón (IIS Aragón), Zaragoza, Spain; Biocomputing Unit, Instituto Aragones de Ciencias de la Salud (IACS), Zaragoza, Spain; Department of Medicine, Psychiatry and Dermatology, University of Zaragoza, Zaragoza, Spain; Endogenous Retroviruses (ERVs) in Development and Disease Group, Instituto Aragones de Ciencias de la Salud (IACS), Zaragoza, Spain; Institut des Maladies Métaboliques et Cardiovasculaires, INSERM, University Toulouse III–Paul Sabatier, UMR 1297-I2MC, Toulouse, France; Research Network on Chronicity, Primary Care and Health Promotion (RICAPPS), Carlos III Health Institute, Madrid, Spain; ARAID Foundation, Zaragoza, Spain

## Abstract

Long COVID (LC) is a broad label encompassing the heterogeneous long-term consequences of SARS-CoV-2 infection that persist for at least 2–3 months post-infection. Despite its substantial global burden, there is still a lack of reliable biomarkers or panels capable of distinguishing individuals with Long COVID from healthy individuals or from those who have recovered from acute COVID-19. We previously characterized a biomarker panel comparing 85 adults with WHO-defined Long COVID against 85 age- and sex-matched controls who had recovered within three months of acute COVID-19 in 2020, at 12–24 months post-infection. That initial profile evaluated blood cell counts, coagulation status, routine SARS-CoV-2 serology, immune cell populations, and basic cytokine levels based on Luminex assays. We have enhanced our biomarker panel by incorporating more precise cytokine quantification using the high-precision ELLA automated immunoassay system. To assess potential ongoing peripheral systemic inflammation, we measured classical inflammatory markers in blood, including C-reactive protein (CRP), tumor necrosis factor alpha (TNFα), interleukin (IL)-1β, and IL-6. In this manuscript, we present data that confirm age- and gender-matching between the LC and control group; and compared differences in cytokine levels and comorbidities.

## INTRODUCTION

Long COVID (LC) is a heterogeneous condition characterized by persistent or fluctuating multi-organ symptoms that can persist for years following a probable or confirmed SARS-CoV-2 infection. This condition comprises a diverse spectrum of partially overlapping symptoms—including fatigue, anosmia, dyspnea, and cognitive dysfunction—whose underlying mechanisms vary and/or remain ill-defined [1, 2]. Since 2023, we have phenotyped a cohort of 85 adults with WHO-defined Long COVID and 85 age- and sex-matched controls who had recovered within three months of acute COVID-19 in 2020 [3]. Unlike most existing research, our study compared Long COVID patients with controls who were infected by the same viral strain at the same time and managed in the same outpatient setting. In a previous study on this cohort [3], we profiled the participants’ immunological, biochemical, and coagulation status, examining a diverse but not exhaustive set of biomarkers. Data confirm the high heterogeneity of LC with respect to clinical manifestations, biomarker profiles, and underlying pathophysiological mechanisms, such as immune dysregulation, viral persistence, and endothelial dysfunction.

Persistent activation of inflammatory pathways [4,5] and increased levels of inflammatory cytokines such as IL-6, CRP, and TNF-α [5-9] have been associated with LC. However, in cohorts recruited from non-hospitalized patients, individuals with LC disease showed no consistent perturbations in standard inflammatory or immunological measurements compared with controls when assessed more than three months post-infection [10, 11]. As we previously found only minor increases in inflammation markers in our cohort [3], we continued phenotyping the cohort focusing on inflammation markers and markers of endothelial damage. High quality measurements were performed on an ELLA automated immunoassay system which includes intraplate normalization. We compared levels of cytokines between LC and non-LC control groups. We also collected comorbidity data for all participants. We hope that this enlarged and improved set of biomarkers will support future efforts at diagnosis and patient stratification.

## MATERIALS AND METHODS

### Study Design and Participants

We analyzed a cross⍰sectional cohort of 170 adults previously described [3], comprising Long COVID (LC; n = 85) and convalescent controls who fully recovered within three months after acute infection (cC; n = 85). Participants were recruited in 2022 from Primary Health Care Centers in Zaragoza (Spain) via routine visits, poster advertisements, and telephone contact after electronic medical record review. Groups were matched by age, sex, and date of acute COVID⍰19 diagnosis. Detailed inclusion/exclusion criteria and diagnostic procedures are provided in the reference publication.

All participants provided written informed consent. The study was approved by the Clinical Research Ethics Committee of Aragón (protocol PI21/278) and conducted in accordance with the Declaration of Helsinki and applicable data protection regulations (EU GDPR 2016/679 and Spanish Organic Law 3/2018). Biological samples and associated data were managed by the Aragón Health System Biobank (National Registry of Biobanks B. B.0000873), under standardized operating procedures and approved governance.

### Data Collection and Processing

Peripheral blood samples were collected and processed for biochemical, coagulation, hematological and flow cytometry analyses as previously described [3].

Levels of the target cytokines were measured using the ELLA automated immunoassay system (Bio-Techne, San Jose, CA, USA). Analyses were performed with a disposable SimplePlex microfluidic cartridge. Prior to loading, serum samples were thawed on ice, diluted in the appropriate sample buffer, and combined with high and low control solutions. Inside the cartridge, each sample was distributed into four separate microfluidic channels, each dedicated to one of the four proteins assessed. Every channel contained three glass nanoreactors specific to the analyte, enabling triplicate measurements for each protein per sample. Cartridges included a lot-specific standard curve for accurate quantification. All assay steps were executed automatically by the instrument without manual intervention. Data were processed by the system software and reported as concentrations in pg/mL.

Comorbidity data for all participants and persistent symptoms for individuals with Long COVID were obtained through standardized clinical questionnaires and coded according to predefined criteria detailed before [3].

### Statistical analysis

Descriptive statistics were computed for all biomarkers (n = 165) across cC and LC. The following metrics were reported: sample size (n); mean and standard deviation (SD); 95% confidence interval (CI) for the mean; median, the interquartile range (IQR); and 95% CI for the median.

For categorical variables, statistical significance was assessed using Chi-square with Yates’ correction when all expected counts were ≥5, or Fisher’s exact test otherwise. Odds ratios (OR) with 95% CIs were estimated applying a continuity correction of +0.5 when any cell contained a zero.

Biomarkers were compared for cC vs LC using Shapiro–Wilk to assess normality. Normally distributed variables were compared with Welch’s t test, and non-parametric variables with Mann–Whitney U test. Effect sizes were reported as Hedges’ g for parametric tests and rank⍰biserial correlation for non-parametric tests. Multiple testing (n=165) across biomarkers was controlled using the Benjamini-Hochberg false discovery rate (FDR) (q < 0.05). Biomarkers with yielded concentrations below the lower limit of detection (LLOD) have not been included in the statistical analyses.

Analyses were performed in R 4.5.1 (dplyr, epitools, stats, ordinal, ggplot2).

As the majority of IL⍰2, IL⍰4, IL⍰17A analyses yielded concentrations below LLOQ, this data was not further analyzed.

## RESULTS

### Study cohort and analytical scope

The present analysis was conducted on a cohort of 170 participants, previously characterized in detail [100]. It comprised two parallel groups: Long COVID (LC, n = 85) and convalescent controls (cC, n = 85), matched by age, sex, and date of acute COVID-19 diagnosis. LC participants were recruited from Primary Health Care Centers in Zaragoza, Spain, during 2022.

Demographic variables, comorbidities, and persistent symptoms were recorded to enable integrated phenotypic characterization.

### Demographic characteristics

LC and cC groups showed comparable demographic profiles, with no meaningful differences in age or sex distribution. Mean age was 48.1 ± 9.7 in cC and 47 ± 10 years in LC, and women represented the majority in both groups (77.7% and 80%, respectively; Figure 1, Table 1).

**Table 1.** Demographic characteristics of study participants in the main groups and subgroups. Summary of age (mean, 95% CI and standard deviation) and sex distribution (n, %) for Long COVID (LC), convalescent Controls (cC), and the C0 and C1 subgroups.

|  | Group | n | Mean | SD | 95% CI of mean |  |
| --- | --- | --- | --- | --- | --- | --- |
|  |  |  |  |  | Lower limit | Upper limit |
| **Age** | cC | 85 | 48.1 | 9.71 | 46 | 50.2 |
|  | LC | 85 | 47 | 10 | 45 | 49 |
| **Gender** |  | **LC (n, %)** | **cC (n, %)** |  |  |  |
|  | Female | 68 (80) | 66 (77.7) |  |  |  |
|  | Male | 17 (20) | 19 (22.3) |  |  |  |

**Figure 1.**
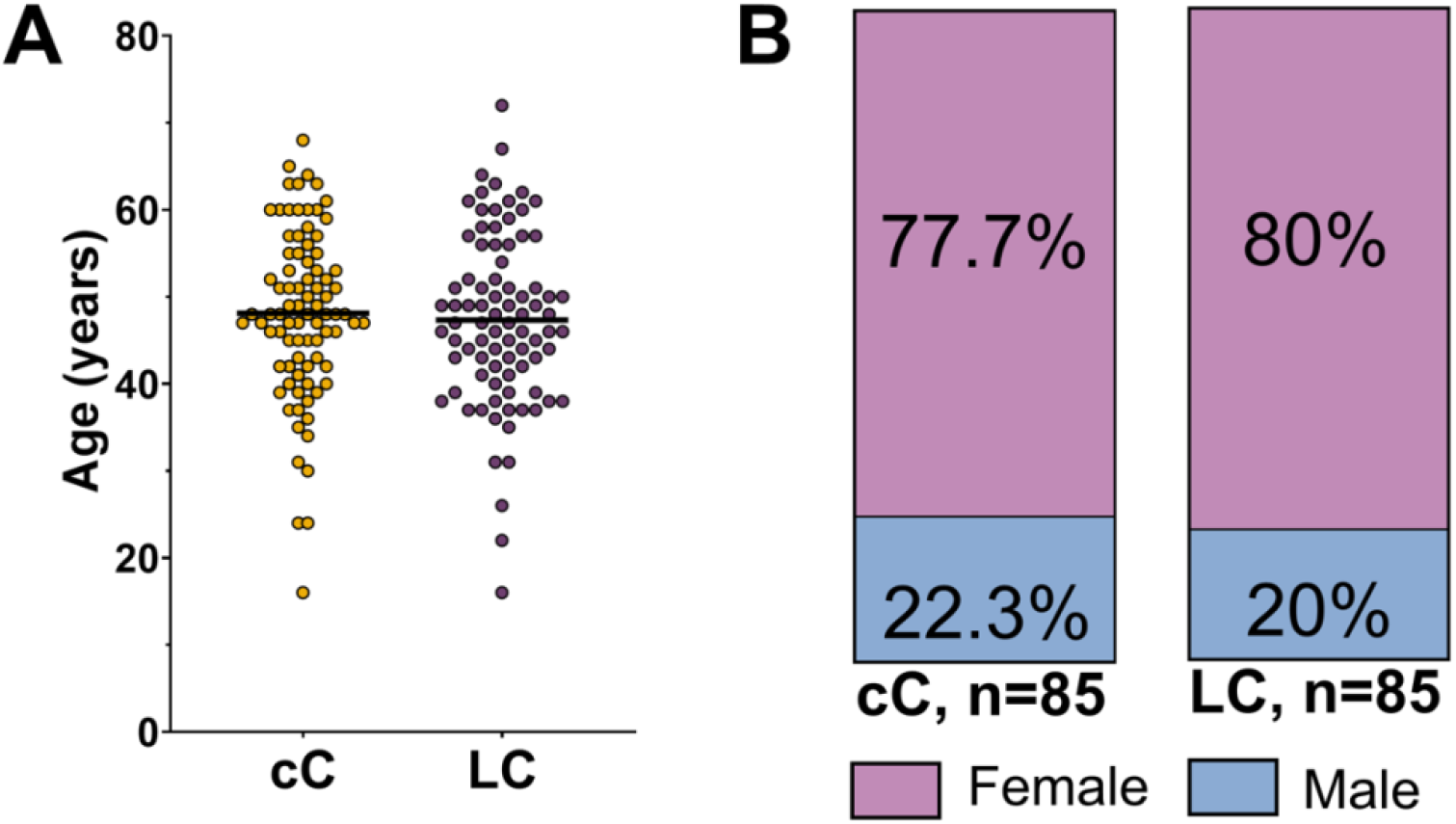
Demographic characteristics of study participants. (A) Age distribution across Long COVID patients (LC), convalescent Controls (cC), cluster C0, and cluster C1. Each dot represents an individual participant; horizontal lines indicate mean values. (B) Sex distribution shown as bar charts for each group, with percentages of male and female participants indicated.

### Comorbidity profiles

Allergies were reported in 22.4% of cC and 55.3% of LC participants, corresponding to a significantly reduced odds ratio in the control group (OR⍰=⍰0.23, 95% CI: 0.12–0.45; p⍰=⍰0.0010). Oral herpes was also more frequent among LC individuals (35.3%) compared to cC (10.6%), and this difference reached statistical significance (OR⍰=⍰0.22, 95% CI: 0.10–0.49; p⍰=⍰0.0127). Additionally, non-specific cardiopulmonary conditions were exclusively observed in the LC group (14.1%), with a markedly low odds ratio for controls (OR⍰=⍰0.03, 95% CI: 0.00–0.59; p⍰=⍰0.0474). Mental health disorders were more prevalent in the LC group (16.5%) compared to cC (n⍰=⍰2), although this difference did not reach statistical significance (OR⍰=⍰0.12, 95% CI: 0.03–0.56; p⍰=⍰0.1853) (Table 2).

**Table 2.** Prevalence of the most frequent comorbidities across convalescent Controls (n = 85), and Long COVID patients (n=85). The table reports the percentages and number of patients that present each comorbidity, along with odds ratios (OR), 95% confidence intervals and FDR-adjusted q-values for pairwise comparisons. Significance was defined as q-value < 0.05. Full comorbidity data across all groups is provided in Supplementary Table S1.

| Comorbidities | n (%) |  | OR | 95% CI |  | p-values |
| --- | --- | --- | --- | --- | --- | --- |
|  | cC | LC |  | Lower limit | Upper limit |  |
| Allergies | 19 (22.4) | 47 (55.3) | 0.23 | 0.12 | 0.45 | 0.0010 |
| Obesity | 14 (16.5) | 24 (28.2) | 0.50 | 0.24 | 1.05 | >0.9999 |
| Oral herpes | 9 (10.6) | 30 (35.3) | 0.22 | 0.10 | 0.49 | 0.0127 |
| Thyroid pathology | 8 (9.4) | 13 (15.3) | 0.58 | 0.23 | 1.47 | >0.9999 |
| Rheumatic and connective tissue disease | 7 (8.2) | 11 (12.9) | 0.60 | 0.22 | 1.64 | >0.9999 |
| Dyslipidemia/<br>hypercholesterolemia | 7 (8.2) | 16 (18.8) | 0.39 | 0.15 | 1.00 | >0.9999 |
| Asthma | 7 (8.2) | 11 (12.9) | 0.60 | 0.22 | 1.64 | >0.9999 |
| Hypertension | 5 (5.9) | 8 (9.4) | 0.60 | 0.19 | 1.92 | >0.9999 |

Other comorbidities, including obesity, thyroid pathology, rheumatic and connective tissue diseases, dyslipidemia/hypercholesterolemia, asthma, and hypertension, were consistently more frequent in the LC group than in controls. However, these differences were not statistically significant.

Overall, while several comorbidities appeared more common among individuals with Long COVID, only allergies, oral herpes, and non-specific cardiopulmonary conditions showed statistically significant associations, suggesting a potential link between these conditions and Long COVID status.

### Biomarkers, cytokines

We next compared circulating cytokines between convalescent controls (cC) and Long COVID participants (LC) to evaluate whether systemic alterations distinguished both groups (Table⍰3).

**Table 3.**
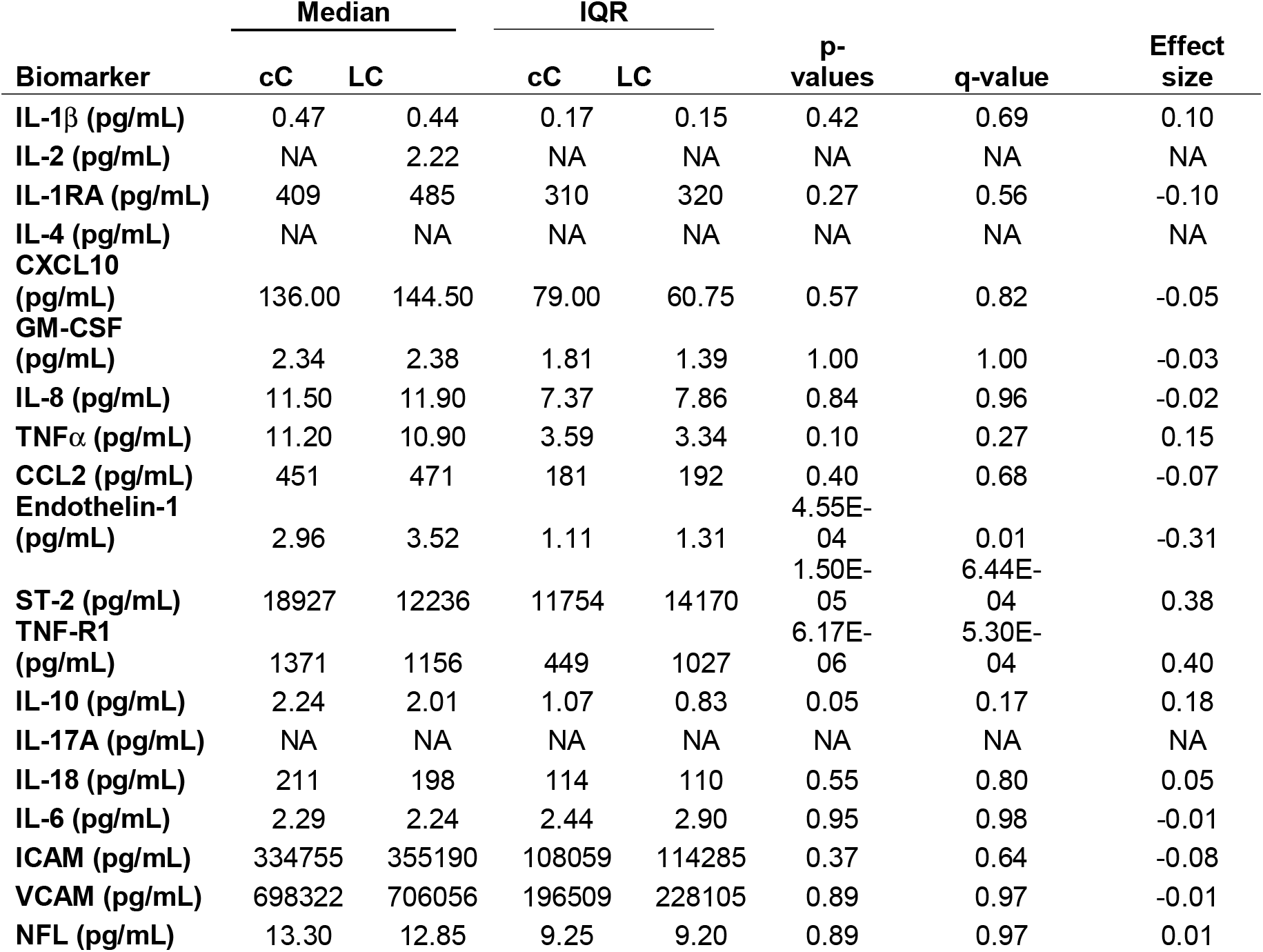
Circulating cytokines. Comparison of circulating cytokines between convalescent controls (cC) and Long COVID (LC) participants. Median concentrations and interquartile ranges (IQR) are shown together with p-values, FDR⍰adjusted q-values, and effect sizes for the cC vs. LC comparison. Normality was assessed using the Shapiro-Wilk test; group differences were examined using Welch’s t-test for normally distributed variables and the Mann-Whitney U test otherwise. Effect sizes correspond to Hedges’ g (parametric) or rank-biserial correlation (non-parametric). Multiple testing was controlled using the Benjamini-Hochberg false discovery rate. Biomarkers with values below the lower limit of detection (LLOD) were excluded from statistical testing. Full biomarker data across all groups is provided in Supplementary Table S2.

After ELLA analysis, concentrations of IL⍰1β, IL⍰1RA, CXCL10, GM⍰CSF, IL⍰8, TNFα, CCL2, IL⍰18, IL⍰6, ICAM and VCAM displayed negligible effect sizes and non⍰significant differences after false discovery rate (FDR) correction (all q⍰>⍰0.17), indicating that baseline inflammatory activity was broadly comparable between groups. Several cytokines (IL⍰2, IL⍰4, IL⍰17a) were below the lower limit of detection in a sufficient number of participants and were therefore not analyzed.

By contrast, three biomarkers showed consistent and statistically significant higher values in LC, even after correction for multiple testing. Endothelin⍰1 concentrations were higher in LC compared with cC (median 2.96 vs 3.52⍰pg/mL in cC; p⍰=⍰4.55×10^− 4^, q⍰=⍰0.01, effect size⍰=⍰–0.31). sST2 and sTNFR1, two markers associated with tissue stress and chronic immune activation also showed robust differences. Levels of sST2 were markedly increased in LC (median 18,927⍰pg/mL vs 12,236⍰pg/mL; p⍰=⍰1.50×10^− 5^, q⍰=⍰6.44×10^− 4^, effect size⍰=⍰0.38). Similarly, sTNFR1 concentrations were significantly higher in LC than in controls (median 1,371 vs 1,156⍰pg/mL; p⍰=⍰6.17×10^− 6^, q⍰=⍰5.30×10^− 4^, effect size⍰=⍰0.40).

Among regulatory cytokines, IL⍰10 exhibited a borderline but non⍰significant difference after FDR correction (q⍰=⍰0.17). Also, markers of neuronal injury (NFL) and endothelial activation (ICAM, VCAM) showed no measurable separation between groups (all q⍰≥ ⍰0.64).

## DISCUSSION

### Strategy

We consider LC a heterogeneous disease, as distinct underlying mechanisms may lead to a similar but also variable disease patterns [4]. Before targeted, individualized therapies can be designed and implemented, a much better understanding of the scope of underlying mechanisms is required.

We have implemented an approach in which data are first collected on an extensive but non-exhaustive set of biomarkers, which will be analyzed later using machine-learning assisted cluster analysis, to identify biomarkers relevant for stratification of patients.

The analysis presented here will be extended in the near future to assess whether either co-morbidities or treatments have contributed to the strength of particular markers and therefore to clustering. This analysis will help identify potential correlations and confounding factors between biomarkers, pre-existing conditions, and treatments. By evaluating these relationships, we aim to improve the accuracy of biomarker selection, ensuring that only relevant associations are considered and minimizing the risk of misleading results. ⍰Apart from gender and age, potential confounding related to treatments received by LC patients, their vaccination status and comorbidities will be considered in future analysis. Using the strategy outlined above, efforts are underway to stratify patients, which may allow for future comparison of patients based on inflammation status.

### Cytokine data

We previously reported data on cytokine expression using Luminex assays [2]. However, this technique is known to exhibit substantial inter- and intra-assay variability, which may arise from differences in assay manufacturers, reagent lot numbers, sample and standard dilution procedures, and operator training [12, 13]. Unless specific conditions related to manufacturer and maintaining consistent methodology are imposed, an over three-fold variability was observed for individual markers. A more recent report addresses there has been limited evaluation of the factors contributing to the inter-laboratory variability of assays run on the Luminex platform [14]. Identifying and minimizing these variances are important for optimal longitudinal and multi-site study design, assay harmonization, scientific rigor, and reproducibility. It was concluded that only when adhering strictly to guidelines for best practices, data with low inter-study variability could be obtained. These guidelines included several conditions (such as assaying all samples as a single batch, centralizing analysis, participating in a quality assurance program, and using paramagnetic-bead kits from a single manufacturer using a standardized protocol) that are hard to implement in any study involving cohorts from different hospitals or recruitment centers.

### Inflammation in LC disease

As long-term immune dysregulation and persistent activation of inflammatory pathways has often been associated with LC disease [6, 7], we assessed the levels of relevant markers including IL⍰1β, CXCL10, TNFα, CCL2, and IL⍰6 in our LC cohort. In comparison to a group of matched patients who contracted COVID disease at the same time as the future LC patients but recovered within 3 months (non_LC controls), we did not observe significantly elevated levels of any of these markers, suggesting that peripheral systemic inflammation may not be a dominant feature of LC in our cohort. We also present data that confirm age- and gender-matching between the LC and control group, reducing the likelihood that these variables confound the observed results.

Similar findings have been reported from other cohorts recruited from primary care, community or outpatient settings. Typically, the majority of long COVID patients in such cohorts never required hospitalization, and showed no consistent perturbations in standard inflammatory or immunological measurements compared with controls when assessed more than three months post-infection [2]. Several community-based and outpatient-enriched cohorts [11; 15; 16] also failed to show elevations of conventional inflammation readouts (CRP, IL-6, ferritin, D-dimer) when compared to matched controls.

We conclude that in our predominantly outpatient long COVID populations systemic inflammation cannot serve as the principal drivers of the phenotype. The implication is that the inflammatory phenotypes does not apply to all patients/cohorts, and mechanistic biomarkers such as sST2 and sTNFR1 are required for classification and stratification. Using the strategy outlined above, efforts are underway to stratify LC patients, which may allow for future comparison of patients independent of inflammation status.

## Data Availability

Original data described previously are publicly available at Zenodo (DOI 10.5281/zenodo.8211331). Additional data reported here will also be made publicly available at Zenodo (DOI 10.5281/zenodo. 21533237) upon publication. This paper does not report original code. Any additional information required to reanalyze the data reported in this paper is available from the lead contact upon reasonable request.

https://doi.org/10.5281/zenodo.21533237

## Declarations

### 1. Ethics approval and consent to participate

This study was approved by the Research Ethics Committee of the Community of Aragon (C.P. - C.I. PI21/410 and C.P. - C.I. PI22/123) and written informed consent was obtained from all participants. The sample collection and treatment were carried out in accordance with approved guidelines.

### 2. Availability of data and materials

Original data described previously [3] are publicly available at Zenodo (DOI 10.5281/zenodo.8211331). Additional data reported here are also publicly available at Zenodo (DOI 10.5281/zenodo.21533237). This paper does not report original code. Any additional information required to reanalyze the data reported in this paper is available from the lead contact upon request.

### 3. Consent for publication

All authors consent to the publication of this manuscript.

## Acknowledgments

We thank the Biocomputing Unit, IACS and the Biobank of the Aragon Health System (PT20/00112) integrated in the Platform ISCIII Biobanks and Biomodels, for their continued support and interest. Thanks to the members of the HERVCOV consortium for their strong commitment and collaborative efforts. We particularly want to acknowledge all the participants for their collaboration in this study. We also thank the Long Covid Aragon Patients Association for their contributions and collaboration in carrying out the study.

