## Supplementary Tables for "High quality analysis of circulating biomarkers reveals no evidence of elevated inflammatory markers in a long COVID cohort recruited at a primary care center"

|  | **n, %** | | | |  | **95% CI** | |  |
| --- | --- | --- | --- | --- | --- | --- | --- | --- |
| **Comorbidities** | **cC** | **%** | **LC** | **%** | **OR** | **Lower limit** | **Upper limit** | **p-values** |
| Allergies | 19 | 22.4 | 47 | 55.3 | 0.233 | 0.12 | 0.453 | 0.0010 |
| Obesity | 14 | 16.5 | 24 | 28.2 | 0.501 | 0.238 | 1.053 | >0.9999 |
| Oral herpes | 9 | 10.6 | 30 | 35.3 | 0.217 | 0.095 | 0.494 | 0.0127 |
| Thyroid pathology | 8 | 9.4 | 13 | 15.3 | 0.575 | 0.225 | 1.469 | >0.9999 |
| Rheumatic and connective tissue disease | 7 | 8.2 | 11 | 12.9 | 0.604 | 0.222 | 1.64 | >0.9999 |
| Hematological disease | 7 | 8.2 | 2 | 2.4 | 3.724 | 0.751 | 18.476 | >0.9999 |
| Dyslipidemia | 7 | 8.2 | 16 | 18.8 | 0.387 | 0.15 | 0.996 | >0.9999 |
| Asthma | 7 | 8.2 | 11 | 12.9 | 0.604 | 0.222 | 1.64 | >0.9999 |
| Tumor/neoplasia without metastasis | 6 | 7.1 | 9 | 10.6 | 0.641 | 0.218 | 1.888 | >0.9999 |
| Hypertension | 5 | 5.9 | 8 | 9.4 | 0.602 | 0.189 | 1.92 | >0.999 |
| Migraine/chronic headaches | 3 | 3.5 | 15 | 17.6 | 0.171 | 0.047 | 0.614 | 0.2932 |
| Autoimmune disease | 3 | 3.5 | 0 | 0.0 | 7.255 | 0.369 | 142.632 | >0.9999 |
| Ocular/vision pathologies | 2 | 2.4 | 4 | 4.7 | 0.488 | 0.087 | 2.738 | >0.9999 |
| Non-neurodegenerative muscle disease | 2 | 2.4 | 6 | 7.1 | 0.317 | 0.062 | 1.619 | >0.9999 |
| Mental health disease | 2 | 2.4 | 14 | 16.5 | 0.122 | 0.027 | 0.556 | 0.1853 |
| Fibromyalgia | 2 | 2.4 | 1 | 1.2 | 2.024 | 0.18 | 22.753 | >0.9999 |
| Diabetes mellitus | 2 | 2.4 | 2 | 2.4 | 1 | 0.138 | 7.268 | >0.9999 |
| Dermatological pathologies | 2 | 2.4 | 0 | 0.0 | 5.12 | 0.242 | 108.252 | >0.9999 |
| Urinary or reproductive system disease | 1 | 1.2 | 8 | 9.4 | 0.115 | 0.014 | 0.937 | >0.9999 |
| Trigeminal neuralgia | 1 | 1.2 | 2 | 2.4 | 0.494 | 0.044 | 5.554 | >0.9999 |
| Peripheral vascular disease (including aortic aneurysm) | 1 | 1.2 | 2 | 2.4 | 0.494 | 0.044 | 5.554 | >0.9999 |
| Osteoporosis | 1 | 1.2 | 3 | 3.5 | 0.325 | 0.033 | 3.193 | >0.9999 |
| Herniated disk | 1 | 1.2 | 1 | 1.2 | 1 | 0.062 | 16.254 | >0.9999 |
| Ear, nose and throat pathologies | 1 | 1.2 | 6 | 7.1 | 0.157 | 0.018 | 1.331 | >0.9999 |
| Diverticulitis | 1 | 1.2 | 0 | 0.0 | 3.036 | 0.122 | 75.576 | >0.9999 |
| Chronic liver disease | 1 | 1.2 | 1 | 1.2 | 1 | 0.062 | 16.254 | >0.9999 |
| Chronic kidney disease | 1 | 1.2 | 1 | 1.2 | 1 | 0.062 | 16.254 | >0.9999 |
| Chronic digestive disease | 1 | 1.2 | 0 | 0.0 | 3.036 | 0.122 | 75.576 | >0.9999 |
| Vertigo/hypoorthostatism | 0 | 0.0 | 5 | 5.9 | 0.086 | 0.005 | 1.573 | >0.9999 |
| Stroke | 0 | 0.0 | 1 | 1.2 | 0.329 | 0.013 | 8.202 | >0.9999 |
| Pancreatic colic | 0 | 0.0 | 1 | 1.2 | 0.329 | 0.013 | 8.202 | >0.9999 |
| Non-specific cardiopulmonary conditions | 0 | 0.0 | 12 | 14.1 | 0.034 | 0.002 | 0.591 | 0.0474 |
| Non-labial herpes or (non-)viral diseases | 0 | 0.0 | 2 | 2.4 | 0.195 | 0.009 | 4.13 | >0.9999 |
| Neuropathic disease | 0 | 0.0 | 1 | 1.2 | 0.329 | 0.013 | 8.202 | >0.9999 |
| Myasthenia gravis | 0 | 0.0 | 1 | 1.2 | 0.329 | 0.013 | 8.202 | >0.9999 |
| Lipid metabolism disorder | 0 | 0.0 | 1 | 1.2 | 0.329 | 0.013 | 8.202 | >0.9999 |
| Irritable colon | 0 | 0.0 | 1 | 1.2 | 0.329 | 0.013 | 8.202 | >0.9999 |
| Hiatal hernia | 0 | 0.0 | 1 | 1.2 | 0.329 | 0.013 | 8.202 | >0.9999 |
| Heart failure | 0 | 0.0 | 1 | 1.2 | 0.329 | 0.013 | 8.202 | >0.9999 |
| Heart attack | 0 | 0.0 | 1 | 1.2 | 0.329 | 0.013 | 8.202 | >0.9999 |
| Gastric atrophy | 0 | 0.0 | 1 | 1.2 | 0.329 | 0.013 | 8.202 | >0.9999 |
| Epilepsy | 0 | 0.0 | 2 | 2.4 | 0.195 | 0.009 | 4.13 | >0.9999 |
| Dysautonomia | 0 | 0.0 | 1 | 1.2 | 0.329 | 0.013 | 8.202 | >0.9999 |
| Cognitive impairment | 0 | 0.0 | 3 | 3.5 | 0.138 | 0.007 | 2.71 | >0.9999 |
| Chronic obstructive pulmonary disease | 0 | 0.0 | 2 | 2.4 | 0.195 | 0.009 | 4.13 | >0.9999 |
| Chronic fatigue | 0 | 0.0 | 1 | 1.2 | 0.329 | 0.013 | 8.202 | >0.9999 |
| Chronic duodenitis | 0 | 0.0 | 1 | 1.2 | 0.329 | 0.013 | 8.202 | >0.9999 |
| Celiac disease | 0 | 0.0 | 2 | 2.4 | 0.195 | 0.009 | 4.13 | >0.9999 |

**Supplementary Table 1. Comparison of comorbidity prevalence across two groups: Long COVID patients (LC, n=85) and convalescent Controls (cC, n=85).** The table reports the percentages and number of patients that present each comorbidity, along with odds ratios (OR), 95% confidence intervals, p-values and FDR-adjusted q-values for pairwise comparisons. Statistical tests were selected based on expected cell frequencies: Chi-square test with Yates correction was applied when all expected frequencies were ≥5; Fisher's exact test was used otherwise. Significance was defined as q-value <0.05.

|  | **Median** | |  | **IQR** | | |  | |  | |  | |  | |  | |
| --- | --- | --- | --- | --- | --- | --- | --- | --- | --- | --- | --- | --- | --- | --- | --- | --- |
| **Biomarker** | **cC** | **LC** |  | **cC** | **LC** |  | | **p-value** | |  | | **q-value** | |  | | **Effect size** |
| A Monocytes (%) | 5.05 | 4.90 |  | 2.90 | 3.80 |  | | 0.95 | |  | | 0.98 | |  | | 0.01 |
| Alanine aminotransferase (ALAT) (U/L) | 19.00 | 17.00 |  | 11.00 | 10.00 |  | | 0.60 | |  | | 0.82 | |  | | 0.05 |
| Albumin (%) | 61.40 | 62.15 |  | 4.70 | 5.35 |  | | 0.09 | |  | | 0.25 | |  | | 0.37 |
| Albumin (g/dL) | 4.40 | 4.50 |  | 0.40 | 0.40 |  | | 0.02 | |  | | 0.08 | |  | | -0.21 |
| Albumin/globulin ratio | 1.58 | 1.54 |  | 0.39 | 0.51 |  | | 0.97 | |  | | 0.99 | |  | | 0.01 |
| Aldolase (U/L) | 3.30 | 2.95 |  | 1.80 | 2.40 |  | | 0.72 | |  | | 0.93 | |  | | 0.03 |
| Alkaline Phosphatase (U/L) | 68.00 | 64.00 |  | 26.00 | 21.00 |  | | 0.45 | |  | | 0.71 | |  | | 0.07 |
| anti GAD (UI/mL) | 5.33 | 5.95 |  | 0.90 | 0.91 |  | | 0.00 | |  | | 0.00 | |  | | -0.47 |
| anti nDNA (UI/mL) | 7.10 | 9.77 |  | 4.97 | 4.97 |  | | 0.12 | |  | | 0.31 | |  | | -0.14 |
| Anticardiolipin IgG (U/mL) | 2.55 | 2.90 |  | 1.95 | 2.30 |  | | 0.39 | |  | | 0.67 | |  | | -0.07 |
| Anticardiolipin IgM (U/mL) | 2.00 | 2.10 |  | 2.00 | 2.43 |  | | 0.31 | |  | | 0.60 | |  | | -0.09 |
| anti-CCP (UI/mL) | 4.59 | 4.59 |  | 0.00 | 0.00 |  | | 0.00 | |  | | 0.04 | |  | | 0.12 |
| Antiendomysium antibodies (U/mL) | 0.00 | 0.00 |  | 0.00 | 0.00 |  | | NA | |  | | NA | |  | | 0.00 |
| Anti-MPO antibodies (MPO) (U/mL) | 3.11 | 3.11 |  | 0.00 | 0.00 |  | | 0.00 | |  | | 0.01 | |  | | 0.13 |
| Antinuclear antibodies (U/mL) | 0.00 | 0.00 |  | 0.00 | 0.00 |  | | 0.76 | |  | | 0.93 | |  | | 0.01 |
| Antiparietal Cell Antibody (ACA) | 0.00 | 0.00 |  | 0.00 | 0.00 |  | | 0.36 | |  | | 0.64 | |  | | 0.04 |
| Antiperoxidase Antibodies (UI/mL) | 6.00 | 2.00 |  | NA | NA |  | | NA | |  | | NA | |  | | NA |
| anti-PR3 (U/mL) | 2.22 | 2.22 |  | 0.00 | 0.00 |  | | 0.01 | |  | | 0.07 | |  | | 0.11 |
| Anti-streptolysin (UI/mL) | 99.99 | 99.99 |  | 68.01 | 48.01 |  | | 0.74 | |  | | 0.93 | |  | | -0.03 |
| Antithrombine III (%) | 96.00 | 104.00 |  | 14.00 | 17.25 |  | | 0.00 | |  | | 0.01 | |  | | -0.33 |
| APTT ratio | 0.96 | 0.98 |  | 0.07 | 0.08 |  | | 0.02 | |  | | 0.10 | |  | | -0.20 |
| Aspartate aminotransferase (ASAT) (U/L) | 22.00 | 21.00 |  | 7.00 | 7.00 |  | | 0.77 | |  | | 0.93 | |  | | 0.03 |
| B Lymphocytes (CD19) (%) | 10.36 | 10.61 |  | 4.23 | 5.41 |  | | 0.79 | |  | | 0.94 | |  | | 0.02 |
| B Lymphocytes (CD19) (cell/mm³) | 0.22 | 0.19 |  | 0.12 | 0.16 |  | | 0.32 | |  | | 0.60 | |  | | 0.09 |
| Basophils (%) | 0.70 | 0.60 |  | 0.30 | 0.50 |  | | 0.23 | |  | | 0.50 | |  | | 0.11 |
| Basophils (10³/μL) | 0.00 | 0.00 |  | 0.10 | 0.10 |  | | 0.92 | |  | | 0.98 | |  | | 0.01 |
| Bilirrubin (mg/dL) | 0.57 | 0.55 |  | 0.26 | 0.28 |  | | 0.28 | |  | | 0.57 | |  | | 0.10 |
| C Monocytes (%) | 88.75 | 88.50 |  | 4.83 | 7.00 |  | | 0.59 | |  | | 0.82 | |  | | 0.05 |
| C1 Complement (mg/dL) | 18.00 | 18.80 |  | 3.10 | 3.40 |  | | 0.08 | |  | | 0.24 | |  | | -0.15 |
| C3 Complement (mg/dL) | 94.80 | 99.70 |  | 21.50 | 25.60 |  | | 0.03 | |  | | 0.12 | |  | | -0.19 |
| C4 Complement (mg/dL) | 20.50 | 24.50 |  | 6.00 | 8.50 |  | | 0.00 | |  | | 0.04 | |  | | -0.25 |
| CCL2 (pg/mL) | 451.00 | 471.00 |  | 181.00 | 192.00 |  | | 0.40 | |  | | 0.68 | |  | | -0.07 |
| CD/CD8 (%) | 2.11 | 2.10 |  | 0.95 | 1.34 |  | | 0.90 | |  | | 0.97 | |  | | -0.01 |
| CD16^-^ CD56^+^/NK^+^NKT (%) | 35.70 | 23.25 |  | 27.00 | 26.45 |  | | 0.03 | |  | | 0.13 | |  | | 0.20 |
| CD16^+^ CD56^-^/CD56^-^ (%) | 5.88 | 5.80 |  | 3.85 | 3.03 |  | | 0.59 | |  | | 0.82 | |  | | 0.05 |
| CD16^+^ CD56^+^/NK^+^NKT (%) | 63.20 | 73.60 |  | 25.30 | 26.80 |  | | 0.02 | |  | | 0.08 | |  | | -0.23 |
| CD16^bright^ CD56^-^/CD56^-^ (%) | 2.21 | 1.57 |  | 1.82 | 1.19 |  | | 0.00 | |  | | 0.00 | |  | | 0.41 |
| CD4 CM/CD (%) | 38.00 | 39.25 |  | 13.55 | 10.13 |  | | 0.56 | |  | | 0.82 | |  | | -0.05 |
| CD4 EM/CD4 (%) | 23.80 | 20.65 |  | 11.85 | 8.70 |  | | 0.08 | |  | | 0.24 | |  | | 0.17 |
| CD4^+^ CD8^low^/lymphocytes (%) | 0.52 | 0.58 |  | 0.60 | 0.79 |  | | 0.41 | |  | | 0.69 | |  | | -0.08 |
| CD56^-^/lymphocytes (%) | 83.60 | 84.80 |  | 7.85 | 7.90 |  | | 0.26 | |  | | 0.54 | |  | | -0.11 |
| CD8 CM/CD8 (%) | 12.95 | 14.20 |  | 6.85 | 10.96 |  | | 0.14 | |  | | 0.34 | |  | | -0.14 |
| CD8 EM/CD8 (%) | 41.20 | 35.83 |  | 19.50 | 18.60 |  | | 0.18 | |  | | 0.42 | |  | | -0.21 |
| CD8^-^ CD4^-^ (%) | 3.00 | 2.80 |  | 2.26 | 2.35 |  | | 0.53 | |  | | 0.78 | |  | | 0.06 |
| CD8 effectors/CD8 | 16.70 | 11.85 |  | 14.15 | 10.13 |  | | 0.00 | |  | | 0.02 | |  | | 0.29 |
| CD8^+^ CD4^low^/lymphocytes (%) | 0.38 | 0.51 |  | 0.40 | 0.45 |  | | 0.01 | |  | | 0.07 | |  | | -0.24 |
| CD8^naive^/CD8 (%) | 22.10 | 29.40 |  | 20.48 | 24.93 |  | | 0.04 | |  | | 0.16 | |  | | -0.19 |
| CD^naive^/CD4 (%) | 33.40 | 36.40 |  | 17.38 | 14.23 |  | | 0.05 | |  | | 0.17 | |  | | -0.18 |
| Cholesterol no-HDL (mg/dL) | 141.00 | 156.00 |  | 49.00 | 46.00 |  | | 0.01 | |  | | 0.04 | |  | | -0.24 |
| CMV2 IgM | 0.00 | 0.00 |  | 0.00 | 0.00 |  | | 0.01 | |  | | 0.04 | |  | | -0.12 |
| C-peptide (ng/mL) | 1.80 | 1.90 |  | 0.89 | 1.07 |  | | 0.32 | |  | | 0.60 | |  | | -0.09 |
| C-reactive protein (CRP) (mg/dL) | 0.17 | 0.17 |  | 0.31 | 0.37 |  | | 0.57 | |  | | 0.82 | |  | | -0.05 |
| Creatine kinase MB (ng/mL) | 1.60 | 1.30 |  | 1.55 | 1.00 |  | | 0.00 | |  | | 0.02 | |  | | 0.28 |
| Creatinine (mg/dL) | 0.70 | 0.78 |  | 0.18 | 0.18 |  | | 0.00 | |  | | 0.02 | |  | | -0.28 |
| Cryoglobulins (%) | 0.00 | 0.00 |  | 1.00 | 2.00 |  | | 0.66 | |  | | 0.88 | |  | | -0.03 |
| CXCL10 (pg/mL) | 136.00 | 144.50 |  | 79.00 | 60.75 |  | | 0.57 | |  | | 0.82 | |  | | -0.05 |
| D-dimer (μg/L) | 271.00 | 292.50 |  | 175.50 | 263.25 |  | | 0.76 | |  | | 0.93 | |  | | -0.03 |
| EBNA IgG | 1.00 | 1.00 |  | 0.00 | 0.00 |  | | 0.74 | |  | | 0.93 | |  | | -0.01 |
| Endothelin-1(pg/mL) | 2.96 | 3.52 |  | 1.11 | 1.31 |  | | 0.00 | |  | | 0.01 | |  | | -0.31 |
| Eosinophils (%) | 2.00 | 2.30 |  | 1.60 | 1.70 |  | | 0.84 | |  | | 0.96 | |  | | -0.02 |
| Eosinophils (10³/μL) | 0.10 | 0.10 |  | 0.10 | 0.10 |  | | 0.88 | |  | | 0.97 | |  | | -0.01 |
| Ferritin (ng/mL) | 87.10 | 74.10 |  | 147.90 | 87.10 |  | | 0.30 | |  | | 0.58 | |  | | 0.09 |
| Fibrinogen (g/L) | 4.10 | 4.40 |  | 1.00 | 1.20 |  | | 0.05 | |  | | 0.16 | |  | | -0.18 |
| Folic acid (ng/mL) | 7.85 | 7.70 |  | 4.95 | 6.20 |  | | 0.72 | |  | | 0.93 | |  | | 0.03 |
| Gamma globulin (%) | 14.40 | 12.80 |  | 2.80 | 2.63 |  | | 0.01 | |  | | 0.05 | |  | | 0.33 |
| Gamma glutamyl transferase (GGT) (U/L) | 21.00 | 20.00 |  | 14.00 | 12.00 |  | | 0.81 | |  | | 0.94 | |  | | -0.02 |
| Globular Sedimentation Rate (GSR) | 9.00 | 11.00 |  | 10.00 | 12.00 |  | | 0.07 | |  | | 0.21 | |  | | -0.16 |
| Glomerular Filtration Rate (mL/min*1.73m²) | 101.82 | 94.26 |  | 15.29 | 18.28 |  | | 0.00 | |  | | 0.02 | |  | | 0.27 |
| Glucose (mg/dL) | 89.00 | 87.00 |  | 14.00 | 11.00 |  | | 0.14 | |  | | 0.34 | |  | | 0.13 |
| Glycosylated hemoglobin (%) | 5.40 | 5.30 |  | 0.40 | 0.50 |  | | 0.02 | |  | | 0.10 | |  | | 0.20 |
| GM-CSF (pg/mL) | 2.34 | 2.38 |  | 1.81 | 1.39 |  | | 1.00 | |  | | 1.00 | |  | | -0.03 |
| Haptoglobin (mg/mL) | 110.00 | 115.00 |  | 64.00 | 69.00 |  | | 0.36 | |  | | 0.64 | |  | | -0.08 |
| HDL (mg/dL) | 62.00 | 62.00 |  | 23.00 | 19.00 |  | | 0.86 | |  | | 0.96 | |  | | -0.02 |
| Hematies (10⁶/μL) | 4.61 | 4.64 |  | 0.61 | 0.46 |  | | 0.91 | |  | | 0.97 | |  | | -0.01 |
| Hematocrite (%) | 40.80 | 41.50 |  | 3.90 | 3.40 |  | | 0.06 | |  | | 0.18 | |  | | -0.17 |
| Hemoglobin (g/dL) | 13.70 | 13.90 |  | 1.30 | 1.10 |  | | 0.25 | |  | | 0.53 | |  | | -0.10 |
| I CAM (pg/mL) | 334755.00 | 355190.00 |  | 108059.00 | 114285.00 |  | | 0.37 | |  | | 0.64 | |  | | -0.08 |
| I Monocytes (%) | 5.85 | 6.00 |  | 3.48 | 3.40 |  | | 0.40 | |  | | 0.68 | |  | | -0.08 |
| Ig A (mg/dL) | 234.00 | 206.00 |  | 128.00 | 149.00 |  | | 0.16 | |  | | 0.37 | |  | | 0.13 |
| Ig D (mg/dL) | 1.76 | 1.58 |  | 4.59 | 3.98 |  | | 0.44 | |  | | 0.70 | |  | | 0.07 |
| Ig G (mg/dL) | 1150.00 | 1040.00 |  | 220.00 | 338.00 |  | | 0.02 | |  | | 0.08 | |  | | 0.21 |
| Ig G Herpesvirus-6 | 1.00 | 1.00 |  | 1.00 | 1.00 |  | | 0.04 | |  | | 0.16 | |  | | -0.16 |
| Ig M (mg/dL) | 116.00 | 118.00 |  | 74.20 | 68.80 |  | | 0.76 | |  | | 0.93 | |  | | -0.03 |
| IgA-tTG (U/mL) | 1.88 | 1.88 |  | 0.00 | 0.00 |  | | 0.72 | |  | | 0.93 | |  | | 0.02 |
| IGFBP-3 (μg/mL) | 5.70 | 6.50 |  | 1.50 | 1.80 |  | | 0.00 | |  | | 0.01 | |  | | 0.59 |
| IgG anti-SARS-CoV2 Spike | 631.00 | 252.00 |  | 1524.75 | 851.00 |  | | 0.07 | |  | | 0.22 | |  | | 0.29 |
| IgG1 immunoproteins (mg/dL) | 529.40 | 475.30 |  | 215.68 | 207.30 |  | | 0.01 | |  | | 0.04 | |  | | 0.25 |
| IgG2 immunoproteins (mg/dL) | 362.40 | 366.30 |  | 151.90 | 190.50 |  | | 0.69 | |  | | 0.92 | |  | | -0.06 |
| IgG3 immunoproteins (mg/dL) | 53.60 | 49.50 |  | 34.63 | 41.90 |  | | 0.12 | |  | | 0.31 | |  | | 0.14 |
| IgG4 immunoproteins (mg/dL) | 29.10 | 28.70 |  | 29.05 | 28.40 |  | | 1.00 | |  | | 1.00 | |  | | 0.00 |
| IL-10 (pg/mL) | 2.24 | 2.01 |  | 1.07 | 0.83 |  | | 0.05 | |  | | 0.17 | |  | | 0.18 |
| IL-17a (pg/mL) | NA | 4.09 |  | NA | NA |  | | NA | |  | | NA | |  | | NA |
| IL-18 (pg/mL) | 211.00 | 198.00 |  | 114.00 | 110.00 |  | | 0.55 | |  | | 0.80 | |  | | 0.05 |
| IL-1b (pg/mL) | 0.47 | 0.44 |  | 0.17 | 0.15 |  | | 0.42 | |  | | 0.69 | |  | | 0.10 |
| IL-1ra (pg/mL) | 409.00 | 485.00 |  | 310.00 | 320.00 |  | | 0.27 | |  | | 0.56 | |  | | -0.10 |
| IL-2 (pg/mL) | NA | 2.22 |  | NA | NA |  | | NA | |  | | NA | |  | | NA |
| IL-4 (pg/mL) | NA | NA |  | NA | NA |  | | NA | |  | | NA | |  | | NA |
| IL-6 (pg/mL) | 2.29 | 2.24 |  | 2.44 | 2.90 |  | | 0.95 | |  | | 0.98 | |  | | -0.01 |
| IL-8 (pg/mL) | 11.50 | 11.90 |  | 7.37 | 7.86 |  | | 0.84 | |  | | 0.96 | |  | | -0.02 |
| International normalized ratio (INR) | 0.93 | 0.93 |  | 0.07 | 0.12 |  | | 0.02 | |  | | 0.09 | |  | | 0.36 |
| Intrinsic Factor Antibodies (AU/mL) | 1.03 | 1.03 |  | 0.08 | 0.06 |  | | 0.05 | |  | | 0.17 | |  | | -0.18 |
| Iron (μg/dL) | 91.00 | 94.00 |  | 42.00 | 35.00 |  | | 0.21 | |  | | 0.46 | |  | | -0.11 |
| Lactate dehydrogenase (LDH) (U/L) | 174.00 | 180.00 |  | 43.00 | 36.00 |  | | 0.46 | |  | | 0.71 | |  | | -0.07 |
| LDL (mg/dL) | 130.00 | 140.00 |  | 40.00 | 47.00 |  | | 0.02 | |  | | 0.09 | |  | | -0.21 |
| Leukocytes (10³/μL) | 6.10 | 6.00 |  | 1.60 | 2.10 |  | | 0.78 | |  | | 0.93 | |  | | -0.03 |
| Lymphocyte total population (%) | 35.00 | 33.60 |  | 13.50 | 13.20 |  | | 0.35 | |  | | 0.64 | |  | | -0.14 |
| Lymphocyte total population (*1000/mm³) | 2.03 | 1.98 |  | 0.70 | 0.77 |  | | 0.27 | |  | | 0.56 | |  | | 0.10 |
| Lymphocytes (10³/μL) | 2.10 | 2.20 |  | 0.80 | 0.80 |  | | 0.81 | |  | | 0.94 | |  | | 0.02 |
| Magnesium (mg/dL) | 2.00 | 2.00 |  | 0.20 | 0.20 |  | | 0.47 | |  | | 0.73 | |  | | -0.06 |
| MCH (pg) | 29.90 | 30.10 |  | 1.90 | 2.00 |  | | 0.51 | |  | | 0.76 | |  | | -0.06 |
| MCHC (g/dL) | 33.70 | 33.50 |  | 0.80 | 1.00 |  | | 0.01 | |  | | 0.04 | |  | | -0.42 |
| MCV (fL) | 88.60 | 90.20 |  | 5.30 | 4.80 |  | | 0.04 | |  | | 0.14 | |  | | -0.19 |
| Medium Platelet Volume (MPV) | 9.10 | 9.10 |  | 1.43 | 1.60 |  | | 0.35 | |  | | 0.64 | |  | | 0.08 |
| Monocytes (%) | 7.80 | 7.50 |  | 2.20 | 2.50 |  | | 0.72 | |  | | 0.93 | |  | | -0.05 |
| Monocytes (10^3^/μL) | 0.50 | 0.50 |  | 0.20 | 0.20 |  | | 0.97 | |  | | 0.99 | |  | | 0.00 |
| Myoglobin (ng/mL) | 21.20 | 20.40 |  | 10.90 | 10.10 |  | | 0.14 | |  | | 0.35 | |  | | 0.13 |
| Neutrophils (%) | 54.50 | 56.10 |  | 14.80 | 12.70 |  | | 0.37 | |  | | 0.64 | |  | | 0.14 |
| Neutrophils (10^3^/μL) | 3.20 | 3.40 |  | 1.80 | 1.60 |  | | 0.74 | |  | | 0.93 | |  | | -0.03 |
| NFL (pg/mL) | 13.30 | 12.85 |  | 9.25 | 9.20 |  | | 0.89 | |  | | 0.97 | |  | | 0.01 |
| NK Lymphocytes (CD56) (%) | 13.04 | 12.60 |  | 8.05 | 6.47 |  | | 0.43 | |  | | 0.69 | |  | | 0.07 |
| NK Lymphocytes (CD56) (cell/m³) | 0.23 | 0.23 |  | 0.21 | 0.13 |  | | 0.29 | |  | | 0.58 | |  | | 0.10 |
| NK/CD56^+^ (%) | 62.80 | 70.05 |  | 15.63 | 28.84 |  | | 0.42 | |  | | 0.69 | |  | | -0.08 |
| NK/lymphocytes (%) | 9.90 | 9.20 |  | 6.33 | 6.39 |  | | 0.65 | |  | | 0.88 | |  | | -0.07 |
| NK^bright^/CD56^+^ (%) | 3.15 | 2.60 |  | 1.94 | 3.01 |  | | 0.37 | |  | | 0.64 | |  | | 0.08 |
| NK^bright^/lymphocytes (%) | 0.44 | 0.41 |  | 0.39 | 0.30 |  | | 0.15 | |  | | 0.36 | |  | | 0.14 |
| NKT/CD56^+^ (%) | 30.80 | 24.88 |  | 21.05 | 28.70 |  | | 0.62 | |  | | 0.85 | |  | | 0.05 |
| NKT/lymphocytes (%) | 4.05 | 3.45 |  | 3.03 | 3.28 |  | | 0.13 | |  | | 0.34 | |  | | 0.14 |
| NT-proBNP (pg/mL) | 45.10 | 45.80 |  | 44.50 | 57.95 |  | | 0.16 | |  | | 0.37 | |  | | -0.13 |
| Partial Thromboplastin Time (TTP) (sec) | 29.80 | 30.30 |  | 2.40 | 2.70 |  | | 0.02 | |  | | 0.10 | |  | | -0.20 |
| Platelets (10^3^/μL) | 242.00 | 248.00 |  | 85.00 | 63.00 |  | | 0.30 | |  | | 0.59 | |  | | -0.09 |
| Potassium (mEq/L) | 4.40 | 4.20 |  | 0.40 | 0.40 |  | | 0.00 | |  | | 0.00 | |  | | -0.60 |
| Prothrombin activity (%) | 120.00 | 121.00 |  | 14.00 | 26.00 |  | | 0.86 | |  | | 0.96 | |  | | -0.03 |
| Prothrombin time (seg) | 11.00 | 11.10 |  | 0.80 | 1.40 |  | | 0.01 | |  | | 0.07 | |  | | 0.38 |
| Red cell distribution width (RDW) (%) | 13.40 | 13.40 |  | 0.90 | 0.90 |  | | 0.93 | |  | | 0.98 | |  | | 0.01 |
| Rheumatoid factor (UI/mL) | 9.99 | 9.99 |  | 0.00 | 0.00 |  | | 0.21 | |  | | 0.47 | |  | | 0.07 |
| Smooth Muscle Antibody (SMA) | 0.00 | 0.00 |  | 0.00 | 0.00 |  | | NA | |  | | NA | |  | | 0.00 |
| Sodium (mEq/L) | 140.00 | 139.00 |  | 2.00 | 3.00 |  | | 0.01 | |  | | 0.04 | |  | | 0.24 |
| sST2 (pg/mL) | 18927.00 | 12236.00 |  | 11754.00 | 14170.00 |  | | 0.00 | |  | | 0.00 | |  | | 0.38 |
| T Lymphocytes (CD3) (%) | 73.53 | 74.36 |  | 10.48 | 8.15 |  | | 0.12 | |  | | 0.31 | |  | | 0.24 |
| T Lymphocytes (CD3) (cell/mm³) | 1.44 | 1.40 |  | 0.63 | 0.65 |  | | 0.67 | |  | | 0.89 | |  | | 0.04 |
| T Lymphocytes (CD4) (%) | 48.34 | 48.15 |  | 11.47 | 11.39 |  | | 0.59 | |  | | 0.82 | |  | | 0.08 |
| T Lymphocytes (CD4) (cell/mm³) | 0.89 | 0.86 |  | 0.46 | 0.43 |  | | 0.47 | |  | | 0.72 | |  | | 0.06 |
| T Lymphocytes (CD8) (%) | 22.46 | 23.69 |  | 8.92 | 11.82 |  | | 0.95 | |  | | 0.98 | |  | | -0.01 |
| T Lymphocytes (CD8) (cell/mm³) | 0.43 | 0.45 |  | 0.21 | 0.34 |  | | 0.79 | |  | | 0.94 | |  | | 0.02 |
| TNF-a (pg/mL) | 11.20 | 10.90 |  | 3.59 | 3.34 |  | | 0.10 | |  | | 0.27 | |  | | 0.15 |
| TNF-R1 (pg/mL) | 1371.00 | 1156.00 |  | 449.00 | 1027.00 |  | | 0.00 | |  | | 0.00 | |  | | 0.40 |
| Total antibodies serology COVID-19 | 1.00 | 1.00 |  | 0.00 | 0.00 |  | | 0.03 | |  | | 0.13 | |  | | 0.12 |
| Total antinucleocapsid antibodies SARS-CoV2 | 1.00 | 1.00 |  | 0.00 | 0.00 |  | | 0.01 | |  | | 0.04 | |  | | 0.14 |
| Total cholesterol (mg/dL) | 205.00 | 228.00 |  | 50.00 | 53.00 |  | | 0.01 | |  | | 0.06 | |  | | -0.23 |
| Total cholesterol/HDL | 3.20 | 3.60 |  | 0.90 | 1.00 |  | | 0.04 | |  | | 0.15 | |  | | -0.18 |
| Total Ig E (UI/mL) | 21.80 | 41.15 |  | 38.70 | 56.03 |  | | 0.01 | |  | | 0.06 | |  | | -0.23 |
| Total protein (g/dL) | 7.00 | 7.10 |  | 0.60 | 0.65 |  | | 0.85 | |  | | 0.96 | |  | | 0.03 |
| Transferrin (mg/mL) | 257.50 | 279.30 |  | 38.70 | 50.60 |  | | 0.01 | |  | | 0.05 | |  | | -0.24 |
| Transferrin saturation (%) | 23.96 | 24.19 |  | 13.44 | 11.99 |  | | 0.80 | |  | | 0.94 | |  | | -0.02 |
| Triglycerides (mg/dL) | 78.00 | 86.00 |  | 54.00 | 55.00 |  | | 0.11 | |  | | 0.29 | |  | | -0.14 |
| TSH (mU/L) | 2.32 | 2.22 |  | 1.24 | 1.49 |  | | 0.98 | |  | | 1.00 | |  | | 0.00 |
| Uric acid (mg/dL) | 4.52 | 4.54 |  | 1.73 | 1.99 |  | | 0.89 | |  | | 0.97 | |  | | -0.02 |
| V CAM (pg/mL) | 698322.00 | 706056.00 |  | 196509.00 | 228105.00 |  | | 0.89 | |  | | 0.97 | |  | | 0.01 |
| Vitamin B12 (pg/mL) | 292.00 | 268.00 |  | 185.00 | 147.25 |  | | 0.10 | |  | | 0.28 | |  | | 0.15 |
| Vitamin D (µg/L) | 57.30 | 70.40 |  | 31.00 | 28.65 |  | | 0.00 | |  | | 0.01 | |  | | -0.30 |
| α-1 Antitriypsin (mg/dL) | 136.00 | 139.00 |  | 21.00 | 23.00 |  | | 0.80 | |  | | 0.94 | |  | | -0.02 |
| α-1 globulin (%) | 3.80 | 3.95 |  | 0.50 | 0.80 |  | | 0.75 | |  | | 0.93 | |  | | -0.04 |
| α-2 globulin (%) | 9.80 | 9.70 |  | 1.90 | 1.58 |  | | 0.96 | |  | | 0.98 | |  | | 0.01 |
| β-2-microglobulin (mg/dL) | 0.17 | 0.18 |  | 0.04 | 0.05 |  | | 0.52 | |  | | 0.77 | |  | | -0.06 |
| β-globulin (%) | 10.80 | 10.65 |  | 1.70 | 1.73 |  | | 0.50 | |  | | 0.75 | |  | | 0.15 |

**Supplemetary Table 2. Comparison of serum biomarkers across clusters cC and LC**. Median concentrations and interquartile ranges (IQR) are reported. P‑values correspond to group comparisons performed using Welch’s t‑test for normally distributed variables and the Mann–Whitney U test otherwise; normality was assessed using the Shapiro–Wilk test. Effect sizes are expressed as Hedges’ g (parametric) or rank‑biserial correlation (non‑parametric). Multiple testing was controlled using the Benjamini–Hochberg false discovery rate, and q‑values are provided accordingly. Biomarkers with concentrations below the lower limit of detection (LLOD) were treated as missing and excluded from statistical testing.
